# Detection of obstetric anal sphincter injuries using machine learning-assisted impedance spectroscopy: a prospective, comparative, multicentre clinical study

**DOI:** 10.1101/2024.06.03.24308379

**Authors:** Katarzyna Borycka, Marcel Młyńczak, Maciej Rosoł, Kacper Korzeniewski, Piotr Iwanowski, Hynek Heřman, Petr Janku, Małgorzata Uchman-Musielak, Erik Dosedla, Enrique Gonzalez Diaz, Iwona Sudoł-Szopińska, Michał Mik, Carlo Ratto, Antonino Spinelli

## Abstract

**Objective:** To evaluate the clinical performance and safety of the ONIRY system for obstetric anal sphincter injuries (OASI) detection versus three-dimensional endoanal ultrasound (EAUS).

**Design:** A prospective, comparative, multicentre, international study. Setting: Poland, Czechia, Slovakia, and Spain.

**Population:** 152 women between the first moments up to 8 weeks after vaginal delivery.

**Methods:** Participants underwent EAUS and were allocated to groups based on OASIS classification: A (no perineal tear), B (1st or 2nd degree tear), or C (3rd or 4th degree, anal sphincters affected). Electric impedance was measured in the anal canal using the ONIRY system. The primary endpoint was the diagnostic outcome of impedance spectroscopy versus EAUS. Adverse events were collected. Part II involved in silico modelling and 10-time 10-fold cross-validation for automated analysis.

**Main Outcome Measures:** Accuracy, sensitivity, and specificity.

**Results:** 30 women were allocated to group A, 61 to group B, and 61 to group C. The diagnostic outcome was determined for 147 participants. The accuracy, sensitivity, and specificity of the ML-assisted impedance spectroscopy were 87.0 ± 0.5%, 90.6 ± 2.0%, and 84.6 ± 1.9%, respectively, compared with EAUS. After data cleaning, the performance metrics of the proposed final ML model for ONIRY were: 90.0 ± 0.4%, 90.0 ± 1.2%, and 90.0 ± 0.7%, respectively. No adverse device effects or deficiencies were observed.

**Conclusion:** ML-assisted impedance spectroscopy offers a safe and accurate method for rapid OASI detection. This approach could effectively complement digital rectal examination in obstetric settings, supporting timely postpartum care.

**Funding information:** The study was financed by the Polish National Centre for Research and Development (POIR.01.01.01-00-0726/18).

**Key Message:** With 90% accuracy, the ONIRY system’s ML-assisted impedance spectroscopy offers a reliable, non-invasive method for detecting obstetric anal sphincter injuries, supporting timely postpartum diagnosis.

## 1. Introduction

Obstetric anal sphincter injuries (OASIs) are common complication of vaginal delivery, directly implicated in various degrees of continence issues, including frank faecal incontinence (FI). It is estimated that one in four women who undergo vaginal delivery experience some form of OASI^1–4^, placing them at a 25-50% risk of developing FI, either shortly after delivery^5,6^ or later in life^7–10^. FI has a profoundly negative impact on quality of life, affecting women’s social interactions, professional activities, family dynamics, and intimate relationships^11–13^.

Despite this significant risk, there is no widely accessible diagnostic tool for early postpartum detection of OASI. Currently, obstetricians rely on digital rectal examination (DRE), the only option in maternity settings, which is highly dependent on the examiner’s experience and skills. As a subjective assessment tool, it is inherently imperfect, with primary detection failure rates for OASI as high as 80%^10^.

The high prevalence of FI among women with undiagnosed OASI^9,10^ underscores the need to revise maternity care protocols. While endoanal ultrasound (EAUS) is the gold standard in the specialise diagnosis of anal sphincter injuries^14,15^, with near-perfect sensitivity in identifying structural abnormalities, it remains limited in terms of availability and usefulness in obstetric facilities. This limitation stems from both a scarcity of qualified staff available in maternity settings to interpret perianal imaging, and the challenging nature of accurate image interpretation in the immediate hours after birth, when the area is often swollen, blood-filled, and secreting fluids^16^, further complicating accurate assessment.

Nonetheless, as highlighted in a Cochrane review^17^, EAUS shows value in the obstetric setting; when used immediately after childbirth, before perineal repair, it can reduce the rate of severe FI at 6-month follow-up. Over the past decade, transperineal ultrasound (TPUS) has shown promise as an alternative to EAUS for identifying OASI, demonstrating good correlation with EAUS results^18,19^. Studies indicate that TPUS, when used immediately postpartum and prior to sphincter repair (if OASI is detected), can identify more OASIs than digital rectal examination alone^20^. However, EAUS still remains the most precise imaging modality available^19^.

To address these clinical gaps, the challenge of developing a novel device - using electric impedance spectroscopy technique has been taken up as the latter is an established physical marker of biostructure/tissue condition^21,22^, already successfully used in other medical applications^23–26^. This technique applies a sinusoidal electrical current below the sensation threshold to the body at various frequencies, measuring the impedance response to infer tissue condition. To date, no known experience exists using impedance spectroscopy for perianal diagnostics. A proof-of-concept study involving 22 patients, followed by two pilot clinical studies using prototype devices on a total of 69 postpartum women, demonstrated the validity of this method^27–30^. Due to subtle differences in impedance values between normal and injured tissue, high sensitivity and specificity were achieved only through the application of nonlinear machine learning (ML) algorithms for test interpretation.

Prototypes of such device, called the ONIRY system, with the ML module gradually trained with the clinical data collected, were designed and developed^27,28^. The overall concept for this system was to serve as a rapid detection method for OASI that could be applied in early postpartum period.

This study aims to evaluate the clinical performance and safety of the ONIRY system—a rapid OASI detection device incorporating impedance spectroscopy and ML—against EAUS as the reference diagnostic method, following re-training of the ML model on a larger, balanced postpartum population.

## 2. Materials and Methods

A prospective, comparative, multicenter, international clinical study was designed, composed of two parts: the clinical conduct (Part I) and modelling and ML (Part II). Part I of the study was conducted from 2021 to 2022 at five European centers in Czech Republic, Slovakia, Poland, and Spain. The study design and conduct were in line with the Good Clinical Practice guidelines for medical device studies (ISO 14155:2020). All approvals by the national regulatory authorities as well as positive opinions by the ethics committees, per local regulations, were obtained prior to study initiation. Written informed consent was collected from each study participant before enrolment. The study was registered at ClinicalTrials.gov under NCT04903977.

### Study design

The study was designed to enroll a total of approximately 150 women between 18 and 49 years old, primiparous, or multiparous, from the first moments up to 8 weeks after vaginal (spontaneous or assisted) delivery of a singleton, live fetus, in any presentation, in gestational week 34 or higher. All inclusion and exclusion criteria are listed in **Supporting Information S1**, along with the detailed study plan.

Three study groups were pre-defined, with fixed numbers of participants enrolled as to ensure generation of balanced data from women with and without OASI. Participants were initially enrolled in these groups based on the evaluation made immediately after delivery according to the 4-degree perineal tears scale^31^: approximately 30 women were planned for group A (no visible perineal tear), approximately 60 for group B (clinically detectable first- or second-degree perineal tear, including episiotomy), and approximately 60 for group C (clinically detectable third- or fourth-degree perineal tear, involving anal sphincters, regardless of primary repair. Specifically, including women with pre-existing primary repairs in Group C was intentional and considered necessary for ML training purposes, as muscle tissue that has been approximated by sutures differs from healthy tissue and should be distinguishable. Furthermore, even after initial sphincter repair, the sphincters’ integrity may remain compromised (in 40-71% of cases) if their continuity is not fully restored^32,33^.

The study duration for each participant was from 2 days up to 5 weeks and included 3 study visits. The first visit, occurring anytime from the immediate postpartum period up to 8 weeks post-delivery, involved the collection of medical history, including pregnancy and birth details, a comprehensive physical examination with proctological and gynaecological assessment, a 12-lead electrocardiogram (ECG) recording, and an evaluation of clinical FI symptoms using the Wexner score. At this initial visit, three dimensional (3-D) EAUS was also performed, serving as the reference diagnostic method and as the final tool for study group allocation.

Following such allocation (per EAUS-based OASIS classification^34,35^), participants underwent impedance spectroscopy using the ONIRY system at the second visit, which took place on the same day as the first visit or up to 7 days later. A web-based application was utilized to provide preliminary test interpretation, allowing the operator to experience an immediate OASI detection result from the ONIRY system (indicating either OASI detected or not detected). These preliminary interpretations were generated from an ML model trained on data from previous pilot studies, which could differ from the final interpretation based on the refined ML model trained during Part II of this study.

To explore the reproducibility of impedance measurements, two consecutive measurement runs were conducted per participant. An arbitrary reproducibility criterion was applied, whereby any difference greater than 1 kΩ at the frequency of 1 kHz between the first and second runs was noted as a discrepancy.

A 12-lead ECG was repeated immediately after impedance measurement with the ONIRY system. Faecal calprotectin levels and blood morphology parameters were also measured to assess any correlation with tissue electrical impedance results. At the third study visit, which occurred between 0 and 28 days after the second visit, anal sphincter function was evaluated using high-resolution anorectal manometry. This procedure was optional, depending on availability at each study site. Vital signs were assessed at each study visit to monitor participant safety throughout. The sequence of study procedures is summarized in Figure 1.

**Figure 1.**
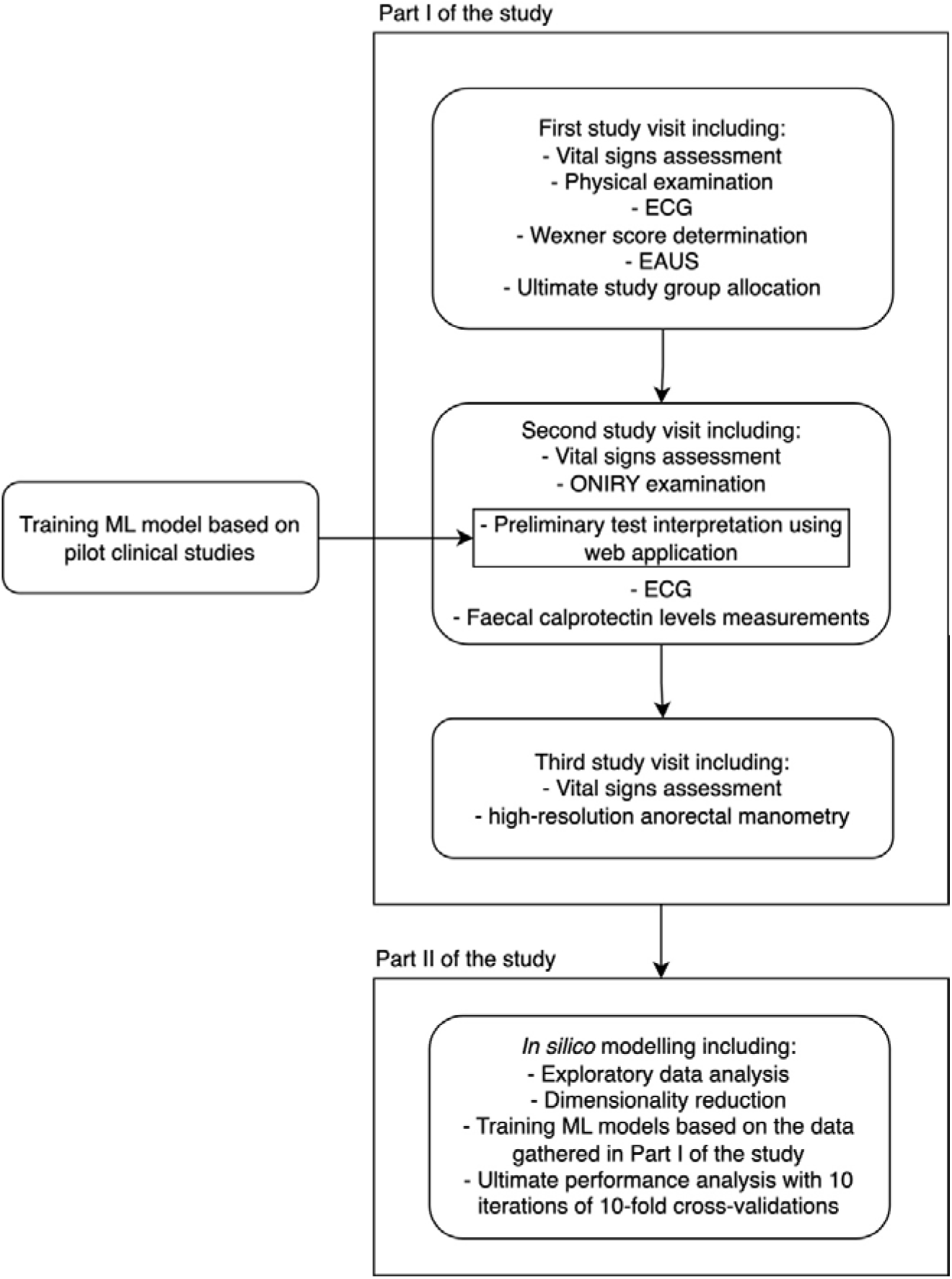
The diagram outlining the individual steps of the study.

Due to the nature of the study design, no blinding was deemed feasible. However, it was not considered essential for maintaining objectivity of study outcomes, as the preliminary interpretation of impedance measurements displayed by the ONIRY system was generated independently based on the ML model trained in prior pilot studies. This interpretation was thus uninfluenced by the test operator and unaffected by knowledge of the EAUS results at study entry. To further minimize any potential bias associated with using EAUS as the primary reference method, a specific technical control measure ensured that the ONIRY examination could only proceed once the EAUS results and interpretation were finalized and entered into the electronic Case Report Form.

#### Study endpoints

The diagnostic outcome of the ONIRY examination compared to 3-D EAUS assessed using the OASIS classification was set as primary endpoint and used for the conclusion on the diagnostic performance of the ONIRY system (following the application of the ML algorithms re-trained in Part II of the study). For the EAUS-based Diagnostic Outcome, OASI was considered detected as long as any depth, length, or circumference range of either anal sphincter (external or internal) was captured (score >2 by OASIS classification). OASI was considered detected if any depth, length, or circumferential involvement of either the external or internal anal sphincter was observed (OASIS classification score >2).

Secondary endpoints related to diagnostic performance assessed in this study, including Diagnostic Outcomes using other reference methods such as digital rectal examination and high-resolution anorectal manometry, were used for respective ML models construction but are not included in this report.

Adverse Events (AE) were recorded for each participant from the time of enrolment until the last study visit.

### Impedance spectroscopy system

Impedance spectroscopy was performed on each study participant using the ONIRY system, which consists of three components: the impedance spectrometer, the endoanal probe, and the ML module. Note that ONIRY is a proprietary name, not an abbreviation. The spectrometer generates a sinusoidal current in the 1-100 kHz frequency range with an amplitude below sensation and pain thresholds, enabling tissue impedance measurement.

The endoanal probe, made from biocompatible, rigid plastic, measures 12 mm in diameter at the electrode site (with a head diameter of up to 19 mm) and contains 8 stainless steel electrodes. These electrodes allow the measurement of impedance modulus, phase shift, resistance, and reactance within the perianal area. The probe also features a handle with a positioning marker to ensure correct placement in the anal canal (see Figure 2).

**Figure 2.**
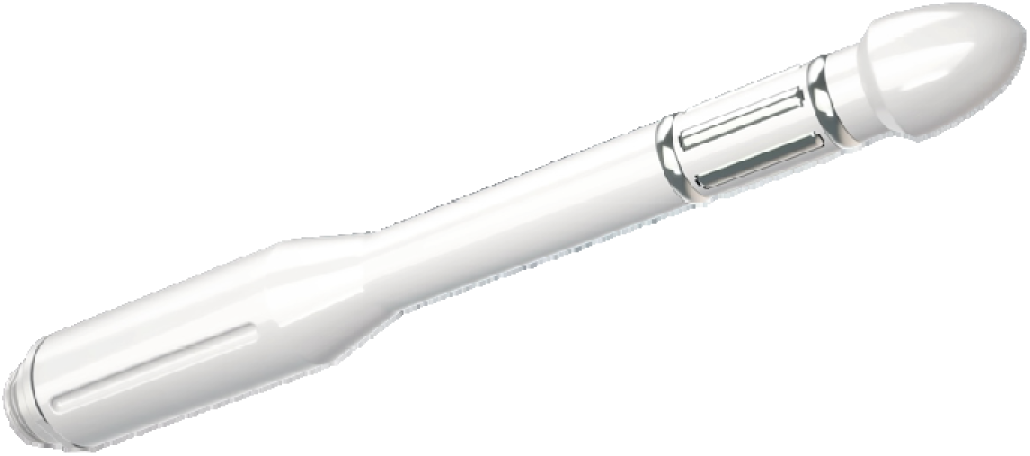
ONIRY Probe with the marker location.

During examination, the probe is inserted into the anal canal for approximately one minute, remaining stationary throughout the measurement. The examination is performed with the patient in a supine position or lying on her left side with knees flexed, depending on operator preference (see Figure 3).

**Figure 3.**
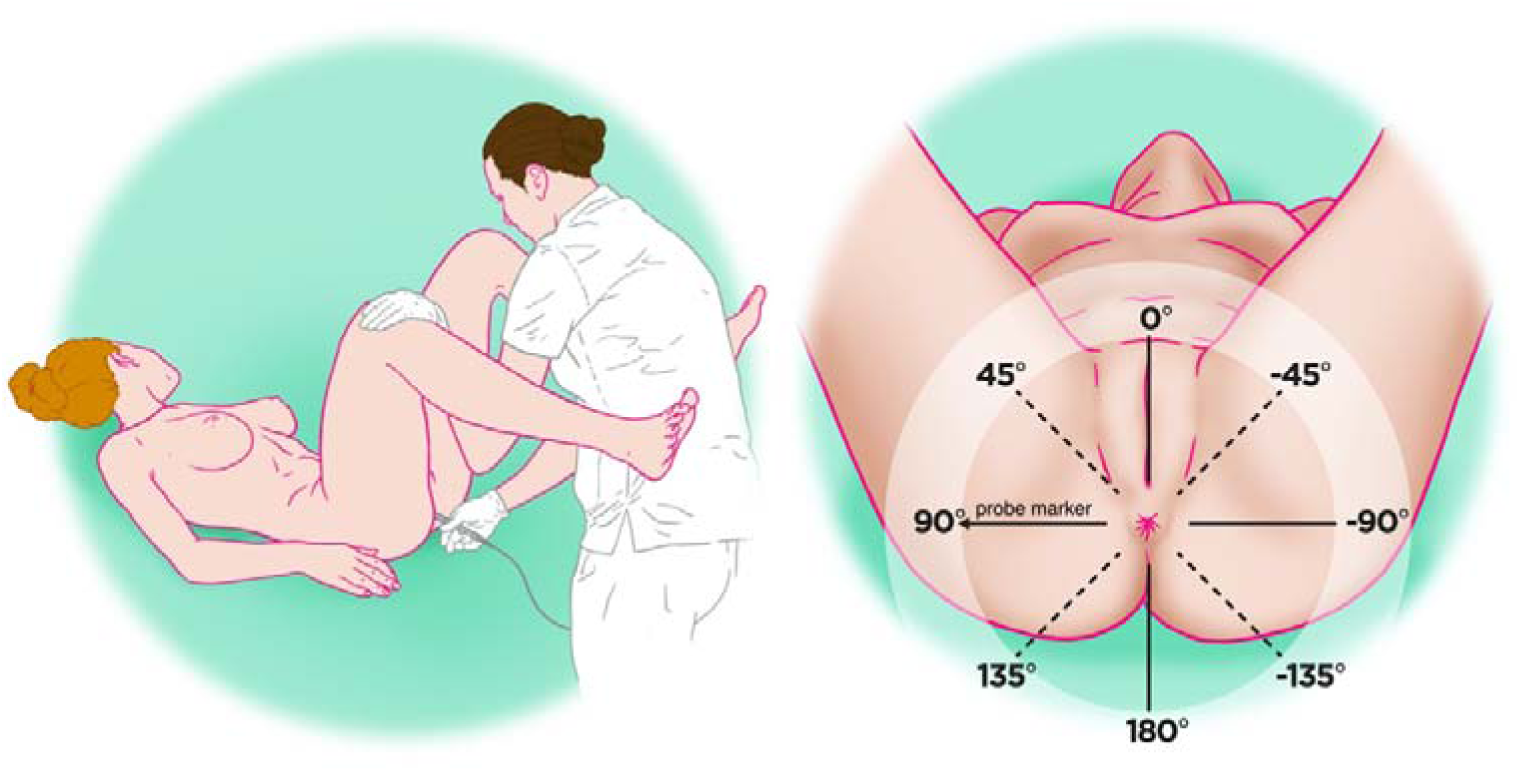
The participant’s position during the ONIRY examination (a) along with position of the probe marker (b).

The spectrometer captures raw impedance data through the endoanal probe, which are then processed to determine statistical parameters for various frequency sub-compartments. These processed parameters serve as an input vector for the ML model, which is trained to analyze subtle differences in impedance patterns that distinguish injured tissue from healthy (or repaired) tissue. The ML model processes these patterns across a complex multi-dimensional dataset, refining its analysis with each frequency parameter to classify tissue integrity accurately.

Following the impedance measurement, the ONIRY system, supported by the ML model, determines whether OASI is present and outputs either “PASS” (no OASI detected) or “REFER” (OASI detected). The ML algorithm is also equipped with control protocols that prevent measurement if improper probe placement is detected, including misalignment or incomplete insertion. The complete ONIRY system setup is illustrated in Figure 4.

**Figure 4.**
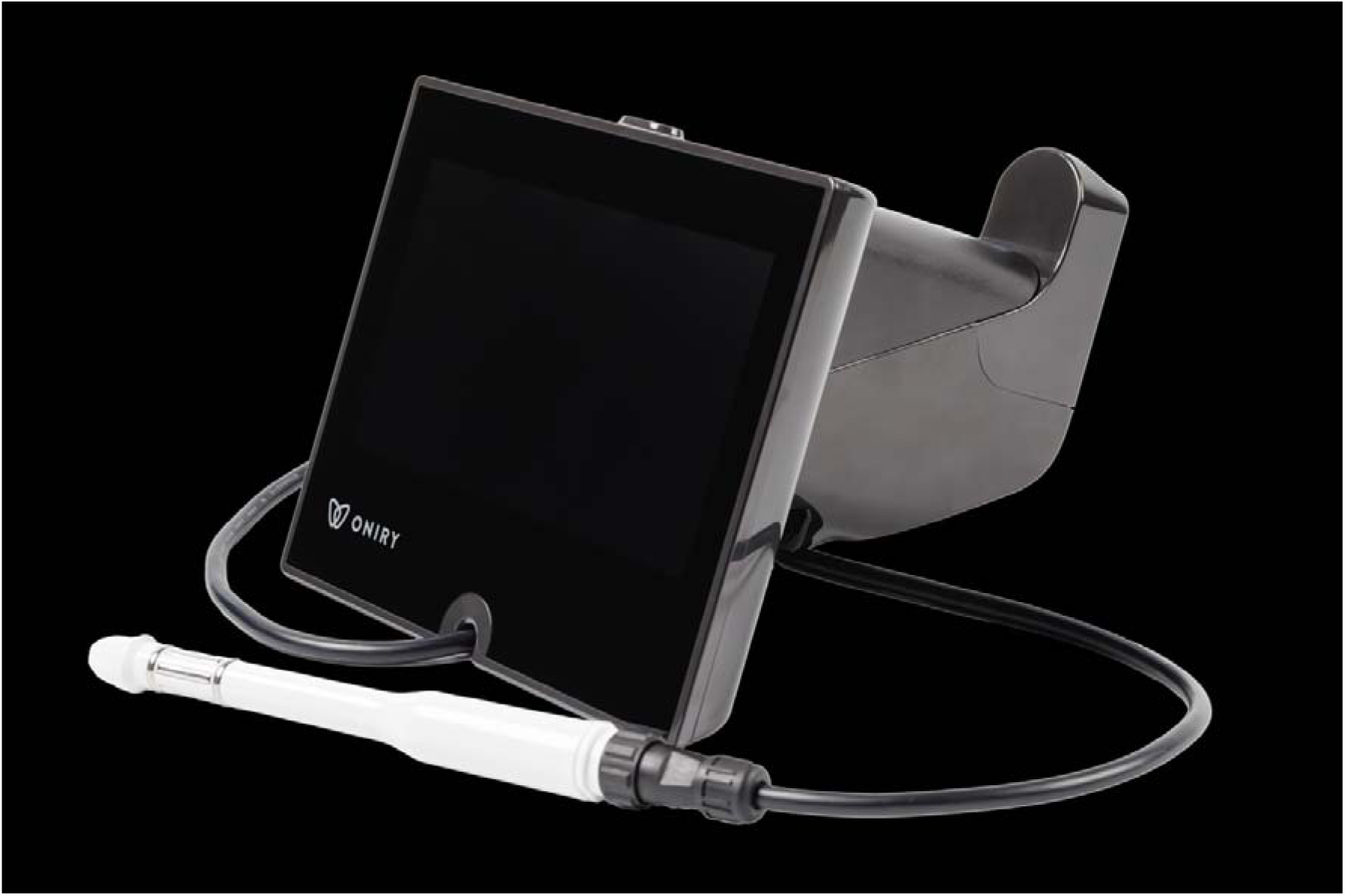
ONIRY system comprising ONIRY Spectrometer and ONIRY Probe.

### Endoanal ultrasound

3-D EAUS, although not typically performed immediately postpartum, remains the only objective, high-precision method for detecting OASI and is thus the gold standard for OASI diagnosis. Despite practical limitations, EAUS was selected as the optimal reference standard for obstetric care in this study to train the ML model and achieve the highest possible diagnostic efficiency for the ONIRY system.

All EAUS examinations were conducted by experts with at least 20 years of experience in perianal imaging. Imaging included assessments of the external and internal anal sphincters as well as the puborectalis muscle. To ensure consistency, the 3-D EAUS procedures followed a standardized study-specific protocol (detailed in a study manual implemented across all centres). The 3-D EAUS results were interpreted according to the OASIS classification^32,33^ for the primary endpoint.

In addition to the OASIS classification, each EAUS examination was assessed using the Starck scale^36^ and the Norderval scale^37^, well-regarded semi-quantitative scoring systems established for EAUS. These alternative assessments allowed for exploratory analyses to determine whether substituting these scores for the OASIS classification could enhance the ONIRY system’s diagnostic performance.

Corresponding ML models were also constructed based on these additional scores, allowing for further evaluation in exploratory studies.

### Statistical analysis

For calculation of diagnostic performance metrics for ONIRY, the Diagnostic Outcome was determined separately for each performance endpoint, as:

1. Diagnostic Success: presence (True Positive) or absence (True Negative) of OASI consistently detected by the ONIRY examination and the reference diagnostic method, or
2. Diagnostic Failure: mismatch (False Positive or False Negative) of the OASI detection by the ONIRY examination and the reference diagnostic method, or
3. Diagnostic Indeterminate: no ONIRY or reference diagnostic method result available or interpretable.

For the primary endpoint, 3-D EAUS result (by OASIS classification) served as the reference method. Evaluation of the exploratory endpoints (diagnostic outcome with 3-D EAUS evaluated with Starck^36^ or Norderval^37^ scales) was performed accordingly. Accuracies were defined as Diagnostic Successes/Total, sensitivities as True Positives/(True positives + False Negatives), and specificities as True Negatives/(True Negatives + False Positives). Also, the F1 score, and Matthew’s Correlation Coefficient (MCC)^38^ were calculated.

The safety profile of ONIRY system was evaluated using descriptive statistics.

### Data analysis (Part I) and Machine Learning modelling (Part II)

Details of the algorithm used to interpret impedance examination results in Part I of the study, based on the preliminary ML model trained with data from two prior pilot clinical studies, are provided in Supporting Information S2. In Part II, in silico analyses included exploratory data analysis, dimensionality reduction, ML modeling, and final performance evaluation with 10-fold cross-validation; full methodological details are available in Supporting Information S3.

For the per-patient reproducibility analysis, if a significant discrepancy was found between the two impedance measurement runs from the same participant, the second measurement run was excluded from the analysis dataset used to develop the final algorithm (note: in real-world application, a single measurement run will be performed per patient).

## 3. Results

### Study population

Of one hundred fifty-three participants screened, 152 were enrolled (1 screening failure). Following 3-D EAUS with OASIS classification, participants were allocated as follows: 30 in Group A (no visible perineal tear and no OASI), 61 in Group B (clinically detectable first- or second-degree perineal tear but no OASI), and 61 in Group C (clinically detectable third- or fourth-degree perineal tear with OASI).

Table 1 presents the key characteristics of the study population by group and overall.

**Table 1.**
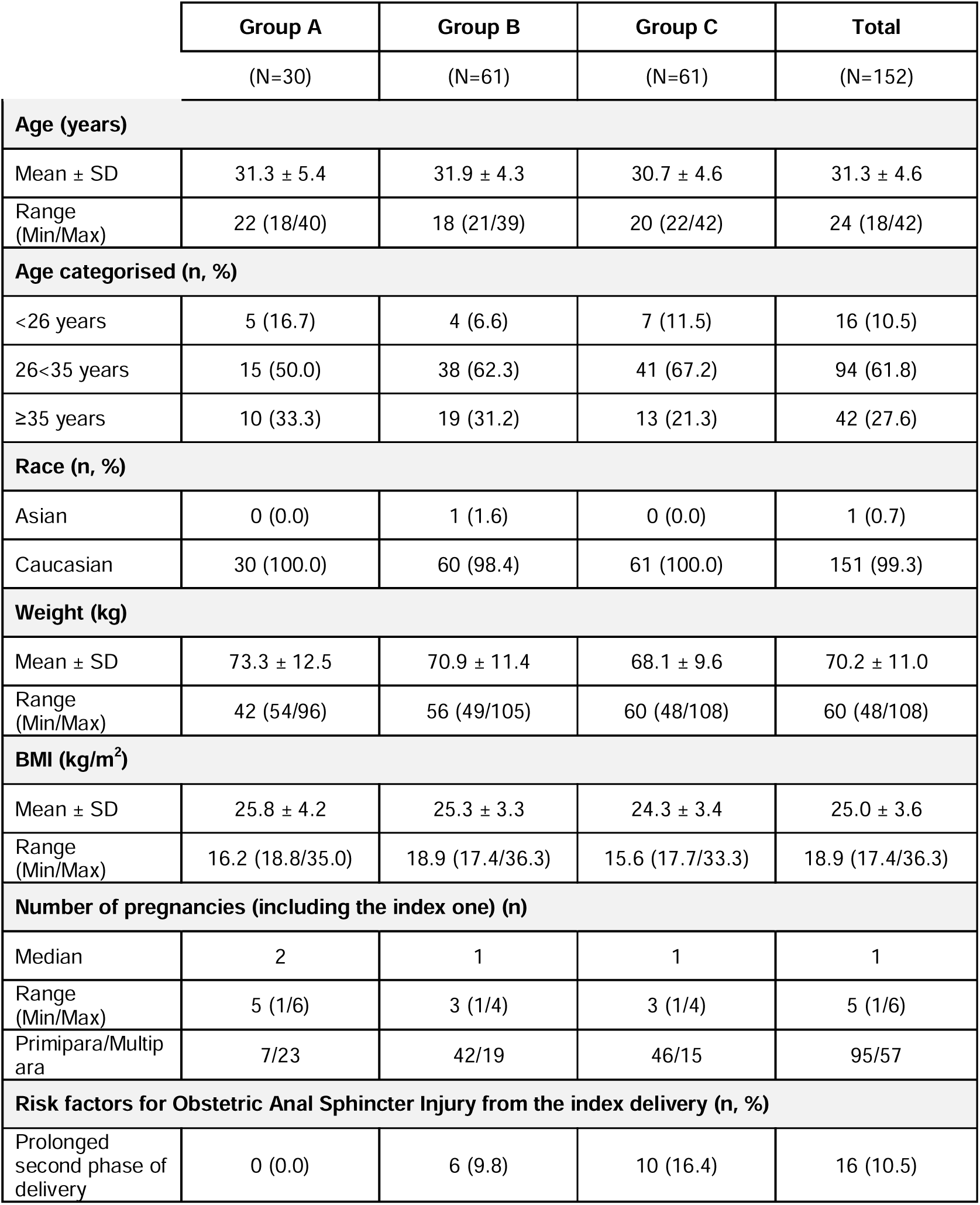
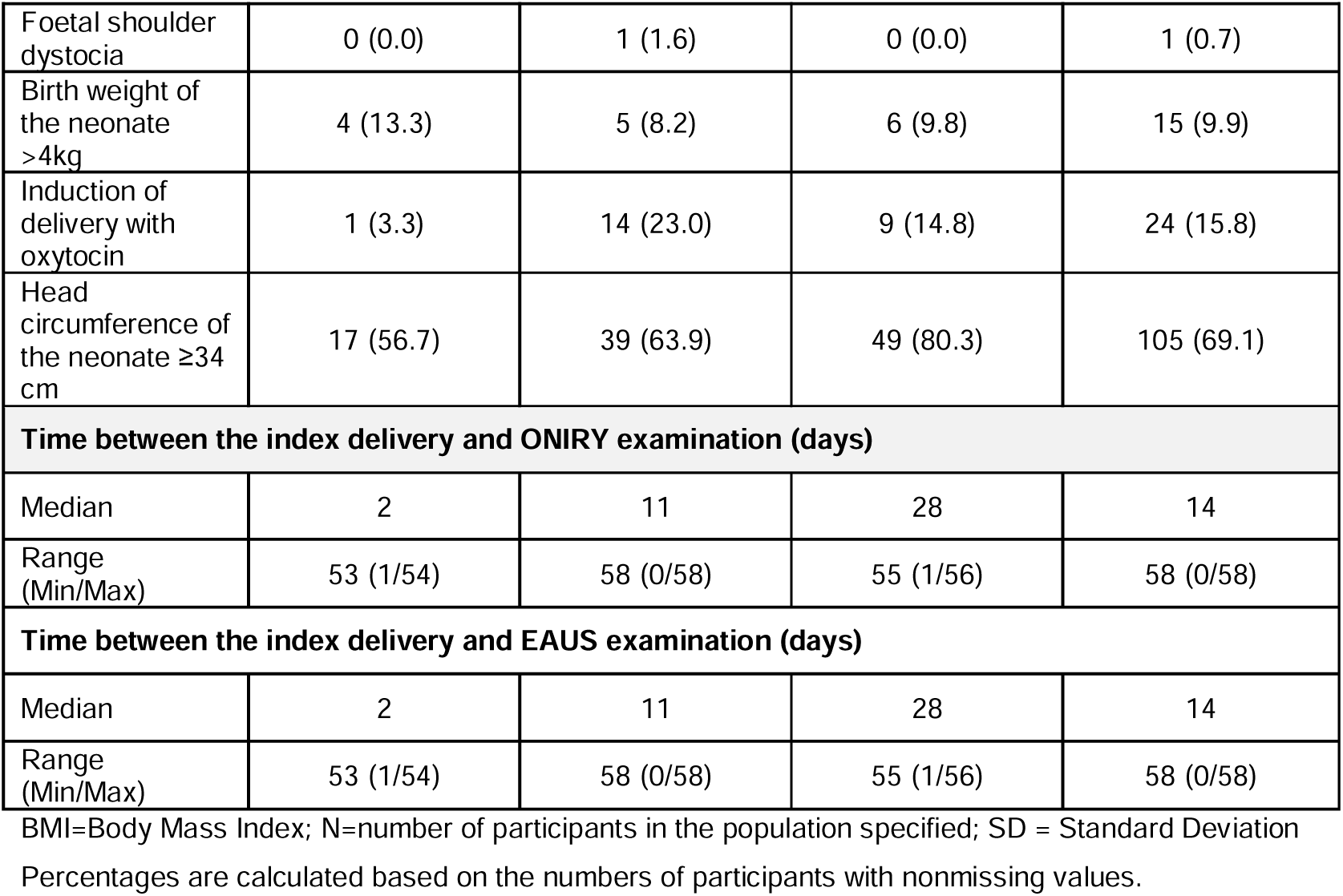
Characteristics of the study population.

Fifteen participants discontinued the study prematurely: 3 in Group A (10.0%; 3/10), 10 in Group B (16.4%; 10/61), and 2 in Group C (3.3%; 2/61), with all discontinuations occurring between the second and last visits. Thus, as all these participants had undergone both EAUS and impedance examinations, they were included in the Clinically Evaluated Population.

### Part I (Clinical conduct)

All enrolled participants were evaluable for the primary endpoint as well as safety outcomes. Diagnostic Success or Failure was determined for 147 participants. For the remaining 5 participants (enrolled at 3 study sites), the Diagnostic Outcome was Indeterminate due to electric impedance measurements falling outside the spectrometer’s expected range, unrelated to the anal canal tissue impedance profile. Thus, the primary endpoint was assessed in 60 participants with OASI (Group C) and 87 participants without OASI (Groups A and B).

Using the preliminary ML model trained on data from prior pilot studies, diagnostic performance metrics were observed as follows: sensitivity of 66.7%, specificity of 57.5%, accuracy of 61.2%, F1 score of 0.58, and MCC of 0.24 (see **Supporting Information S4** for detailed metrics).

No significant correlation was found between faecal calprotectin levels and the results of wither EAUS or ONIRY examinations.

### Part II (*In silico* Machine Learning modelling)

The ultimate performance analysis was conducted with an ML model trained on data generated in Part I of this study, incorporating both raw impedance data from ONIRY and OASI detection results from EAUS as a reference standard. A total of 298 impedance measurements meeting quality requirements (see **Supporting Information S5**) were included: 122 from participants with OASI (group C) and 176 from participants without OASI (group A and B). Following exploratory data analysis, dimensionality reduction, and modelling using artificial neural networks, diagnostic metrics showed marked improvement, achieving sensitivity slightly above 90% and specificity slightly below 85%, as presented in Table 2 (with individual cross-validation statistics available in **Supporting Information S6**).

**Table 2.** Mean performance metrics of the ONIRY system using the models trained on data from Part I of the study, in the assessment relative to 3-D Endoanal Ultrasound and OASIS classification.

|  | **Accuracy [%]** | **Sensitivity [%]** | **Specificity [%]** | **F1** | **MCC** |
| --- | --- | --- | --- | --- | --- |
| **Mean ± SD** | **87.0 ± 0.5** | **90.6 ± 2.0** | **84.6 ± 1.9** | **0.85 ± 0.004** | **0.75 ± 0.01** |

MCC = Matthew's Correlation Coefficient; SD = Standard Deviation

The exploratory per-patient reproducibility analysis from Part I revealed discrepancies between the two impedance measurement runs in 19 participants (12.8%). In exploratory analyses using alternative EAUS classification scores, performance metrics based on the Starck^36^ and Norderval^37^ classifications were assessed. Using the Starck classification sensitivity was 83.8%, specificity 88.1%, accuracy 86.4 %, F1 score 0.83, and MCC 0.73. For the Norderval classification, sensitivity reached 84.3%, specificity 89.3 %, accuracy 87.4 %, F1 score 0.84, and MCC 0.74.

### Safety results

No deaths, serious AE, or AE leading to premature withdrawal from the study were reported during the study conduct. A total of 22 AE were observed in 21 participants: 4 AE in group A, 5 in group B, and 13 in group C. All AE occurred after the ONIRY examinations. Four types of AE were reported by more than one participant, with the most common being nasopharyngitis (2.6%, 4/152) and COVID-19 (2.6%, 4/152).

None of AE were considered related to the ONIRY system, and no adverse device effect were reported.

No trends were observed in ECG parameters when comparing baseline recordings (pre-ONIRY examination) with post-ONIRY recordings. Additionally, no clinically significant cardiovascular abnormalities were recorded as an AE following ONIRY application.

### Post-hoc analysis

Following exploratory analyses of per-patient reproducibility in impedance measurements, 19 measurement files (each from the second measurement per relevant patient) were excluded from the final ML model for the ONIRY system, resulting in a “limited dataset” comprising 93.6% (279/298) of the original dataset. The limited dataset included 279 impedance measurements: 117 corresponding to OASI and 162 with no OASI. A post-hoc performance analysis using cross-validations on this refined ML model demonstrated diagnostic metrics with both sensitivity and specificity of ONIRY system at 90.0%, as detailed in Table 3 (individual cross-validation statistics are provided in **Supporting Information S6**).

**Table 3.** Mean performance metrics of the ONIRY system using the Machine Learning model trained based on the limited clinical study dataset in the assessment relative to 3-D Endoanal Ultrasound and OASIS classification.

|  | **Accuracy [%]** | **Sensitivity [%]** | **Specificity [%]** | **F1** | **MCC** |
| --- | --- | --- | --- | --- | --- |
| **Mean ± SD** | **90.0 ± 0.4** | **90.0 ± 1.2** | **90.0 ± 0.7** | **0.88 ± 0.01** | **0.80 ± 0.01** |

MCC = Matthew's Correlation Coefficient; SD = Standard Deviation

## 4. Discussion

### Key Findings

The ML-supported impedance spectroscopy using ONIRY system demonstrated high diagnostic performance for detecting OASI when compared to EAUS (per OASIS classification) in the study population enriched with OASI cases (40.1%, 61/152 of enrolled women). Following re-training of the ML model in Part II with data generated in Part I, the system achieved an accuracy of 87.0 ± 0.5%, with a sensitivity of 90.6 ± 2.0%, and specificity of 84.6% ± 1.9%.

After refining the dataset by excluding 6.4% (19/298) of impedance measurements due to measurement discrepancies, the optimized ML model intended for the final ONIRY system reached an accuracy of 90.0 ± 0.4%, with both sensitivity and specificity at 90% (sensitivity 90.0 ± 1.2%, specificity 90.0 ± 0.7%). This substantial diagnostic improvement highlights the benefit of ML-supported analysis of impedance data for OASI detection, achieved through re-training with a larger, well-balanced dataset (OASI vs. no OASI) compared to initial pilot studies.

The intermediary performance observed in Part I (accuracy of 61.2%), which used an ML model trained only on smaller, less balanced pilot datasets (with fewer OASI cases)^27–30^, underscores the critical role of dataset size and balance in enhancing diagnostic accuracy. The improved performance of the final ML model in this study likely reflects the broader variability of raw impedance data collected and the higher representation of OASI cases, which better supported model optimization.

Exploratory analyses using alternative EAUS reference methods (Starck and Norderval classifications with corresponding alternative ML models) did not demonstrate additional diagnostic advantages over the primary performance metrics. No safety concerns were identified in association with the ONIRY system.

### Clinical Implications

Currently, no rapid easy-to-use diagnostic tool is available in maternity care settings for whole obstetric team, beyond digital rectal examination. Although DRE is a standard procedure, present in most obstetric guidelines^39–43^, it has significant limitations due to its subjective nature with sensitivity for detecting OASI heavily dependent on the examiner’s experience^44–46^. Hence the crucial importance of practical training and programmes dedicated to midwives and obstetricians aimed at increasing the OASI detection rate and the effectiveness of its management^47,48^.

Although EAUS, as the gold standard for detecting OASI, is rarely feasible in the early postpartum period due to resource and operational constraints, as it requires expert handling and is challenging to interpret images in the immediate postpartum hours^49^ - its value in accurately detecting even minor injuries is undeniable. Typically, EAUS is more suitable later in the postpartum period, around 6-8 weeks after delivery, when patients return with symptoms of incontinence or perineal wound healing issues, which are current indications for an EAUS assessment.

As demonstrated in this study, the ML-supported impedance spectroscopy, providing a straightforward interpretation of the perianal tissue impedance results, has shown high diagnostic accuracy, achieving approximately 90% sensitivity and specificity (compared with 3-D EAUS, per OASIS classification, the only available objective reference) during the whole postpartum period (from the first hours up to 8 weeks after delivery). This level of performance suggests that the impedance spectroscopy using ONIRY could serve as an effective adjunct to DRE, particularly in settings where EAUS is unavailable or limited for initial detection. Since a single measurement run of the impedance (including the automated analysis and presentation of the results) takes less than a minute, it may have an acceptance potential for obstetric practice. The use of such diagnostic tool within the first 24 hours after delivery, could provide the greatest clinical benefit by enabling timely primary repair of OASI, thereby reducing long-term complications.

A systematic review by Walsh et al.^17^ suggests, that performing EAUS immediately after delivery, before any perineal repair, significantly increases OASI detection rates and consequently the rate of primary sphincter repair. Additionally, EAUS conducted prior to perineal repair may reduce the risk of developing severe faecal incontinence. This finding highlights a critical gap in routine obstetric practice, where the absence of efficient tools for OASI detection may prevent timely intervention.

Moreover, impedance spectroscopy could be valuable also in the following weeks postpartum. By bridging the gap until EAUS becomes feasible, the use of ONIRY could expand diagnostic capabilities to include asymptomatic and occult OASI cases that would otherwise go undetected until FI symptoms appear. Proper diagnosis within this time window, even if too late for primary repair, could allow for the identification of patients who require ongoing monitoring or targeted rehabilitation for promoting quicker recovery and minimizing the risk of FI.

What favours EAUS as the diagnostic method is the possibility to separately visualise both external and internal anal sphincters. This enables targeted repairs of the external sphincter alone or combined repair of both sphincters, making EAUS an essential tool for elective diagnosis before any delayed sphincter repair.

Therefore, there is an unmet need for establishing an effective, noninvasive or minimally invasive method of detecting OASI for maternity care facilities and midwifery practices, where reliable diagnostic tools are often unavailable. Ideally, such a method would function as a screening tool with clear criteria for extended diagnosis, tailored to the specific needs and resources of the facility. If impedance spectroscopy proves successful in clinical practice, it could not only enhance postpartum management in maternity care settings but also facilitate timely referrals to specialist surgical units, ultimately improving long-term outcomes for women affected by OASI. Because ONIRY provides automated, rapid interpretation of impedance data in a simple binary output (PASS/REFER), it has potential for routine use by a wider range of maternity care staff, even those with minimal specialized training.

No risks or safety concerns have been identified for the electric impedance-based method for OASI detection, in particular with the use of the ONIRY system. From the performance standpoint, the following contraindications for ONIRY examination could be considered: any implants in the pelvic area, major malformations of the perianal area, or coincident connection with any electronic medical or surgical equipment or device generating alternating current above 1 kHz.

This study represents the first evaluation of impedance spectroscopy for OASI detection in a broader postpartum population. Future studies are planned, focusing on the critical time window within the first few hours postpartum, to further refine the ML model’s efficacy at a point when early OASI detection offers the most benefit— allowing for timely primary repair.

## Limitations and research implications

This study has several limitations.

Firstly, the study population was selectively enriched with OASI cases (Group C, n=61) to improve the dataset’s internal balance and increase the robustness of the ML model. This approach, common in AI and ML tool development^50^, ensures higher internal data coherence, enhancing the model’s effectiveness within a controlled dataset structure.

Secondly, the primary endpoint focused on OASI detection rather than a clinically meaningful outcome, such as faecal incontinence (FI) or quality of life measures. The surrogate endpoint was chosen to facilitate a clear comparison between the ONIRY system and 3-D EAUS, the gold standard, within a manageable timeframe. Establishing a clinically meaningful benefit for the ONIRY system would require extended, long-term follow-up studies, spanning years to account for the gradual development of FI in women with OASI. The potential benefit of rapid OASI detection hinges on the availability and prompt implementation of therapeutic measures, notably primary sphincter repair within the optimal 8–12 hour window after delivery^34,35, 39–43^.

Another limitation arises from the high proportion of participants in Group C (52 out of 61) who had already undergone primary sphincter repair before study enrolment. This factor may have influenced impedance spectroscopy results toward readings typical of non-injured cases. However, given that the ML models were trained with this dataset, it is plausible that the ONIRY system’s performance could be even greater in real-world use, where OASI detection would ideally occur before repair. Newly injured, unrepaired sphincters are likely to present a clearer contrast in impedance measurements compared to tissues that have already undergone repair. The difference in median time between delivery and ONIRY examination across groups (28 days in Group C, versus 2 and 11 days in Groups A and B, respectively) reflects challenges encountered in recruiting cases of 3rd and 4th degree perineal tears (Group C) in this challenging study design. This delay also mirrors the lengthy diagnostic and therapeutic pathway typically experienced by OASI patients in current practice.

In addition, the inclusion of participants with previously repaired OASI in Group C captures clinical scenarios where underdiagnosed or inadequately repaired OASIs persist despite initial repair. Literature indicates that primary repair is incomplete in over 30% of cases, often due to the limited experience of the operator or the emergency nature of the procedure itself, usually performed during on-call hours^51–53^. Thus, classifying cases with recent repairs in the “injured” group is methodologically sound and aligns with real-world conditions.

Finally, no detailed data were captured for multiparous women on potentially undetected OASIs from prior deliveries. This could theoretically result in normal impedance readings due to tissue healing, despite persistent EAUS abnormalities. Conversely, abnormal impedance with normal EAUS could indicate fibrosis from a prior injury.

## 5. Conclusions

The ML-assisted impedance spectroscopy demonstrated safety and high diagnostic accuracy, achieving approximately 90% sensitivity and specificity in detecting obstetric anal sphincter injuries in women after vaginal birth. This approach could effectively complement digital rectal examination in obstetric settings, supporting timely postpartum care.

## Data Availability

Data sharing is not applicable to this article due to legal and privacy
issues.

## Ethics Statement

The study was approved by the ethics committees respective for each study site: on 19 March 2021 by Ethics Committee of the Institute for Maternal and Child Care (no. 1/19.03.2021), on 27 April 2021 by Ethics Committee for Research with Medicines of the Health Areas of León and Bierzo (no. 2186), on 9 June 2021 by Ethics Committee of the University Hospital of Brno (no. 47/21Zdrav.), on 14 October 2021 by Ethics Committee at the Regional Medical Chamber in Warsaw (no. KB/1362/21) and on 25 July 2022 by Ethics Committee at AGEL Hospital Košice-Šaca (no. ONIRY 3/2/2020).

## Conflict of Interest statement

K.B. is a founder and board member at OASIS Diagnostics, an author of the related patent and R&D strategy, independent consultant, and trainer of Takeda.

M.M., M.R., K.K., and P.I. are researchers at OASIS Diagnostics.

A.S. is an independent consultant of Ethicon, Takeda, Pfizer, Sofar. M.UM. is an independent consultant of Regen Lab.

H.H., P.J., M.UM., E.D., and E.GD. received remuneration as a study investigator. H.H., C.R., and A.S. are independent consultants and members of the OASIS Diagnostics’ Scientific Advisory Board.

Others declare no conflicts of interest.

## Funding

The study was funded by the European Union as part of the Fast Track program, conducted in Poland by the National Centre for Research and Development (POIR.01.00.01-00-0726/18).

## Abbreviations

AE: Adverse Events
EAUS: Endoanal Ultrasound
FI: Faecal Incontinence
MCC: Matthew’s Correlation Coefficient
ML: Machine Learning
OASIs: Obstetric Anal Sphincter Injuries

## Author contributions

Conceptualization: KB, MM, PI

Data curation: KB, MM, MR, KK, PI

Formal analysis: MM, MR, KK

Funding acquisition: KB, MM

Investigation: HH, PJ, MUM, ED, EGD

Methodology: KB, MM, MR, KK, PI

Project administration: KB, MM

Resources: HH, PJ, MUM, ED, EGD

Software: MM, MR, KK

Supervision: KB, MM, PI, CR, AS

Validation: MM, PI

Visualization: MR, KK

Writing - original draft: KB, MM, MR, KK, PI

Writing - review & editing: All co-authors

## Supporting information

S1 – Schedule of assessments of the pivotal study (Part I – clinical conduct) valid at the time of the study completion (following Amendment no. 2 of the study protocol).

S2 - The description of the heuristics used for Part I of the study

S3 - Details of the in silico analyses conducted within Part II of the study.

S4 - Performance metrics of the ONIRY model trained based on data from the previous (pilot) clinical studies, depending on the heuristics chosen, with the 3-D Endoanal Ultrasound and the OASIS classification (>2 vs. ≤2) used as the reference method.

S5 - The quality requirements for the impedance measurements.

S6 - Performance metrics of the ONIRY system using the models trained based on all correct measurements from the clinical study, in the assessment relative to 3-D Endoanal Ultrasound and OASIS classification (each row shows the statistics for a single 10-fold cross-validation).

S7 - Performance metrics of the ONIRY system using the model trained based on the limited clinical study dataset (discrepant measurements, considered based on the arbitrary criterion set in Statistical Analysis Plan, were ignored), in the assessment relative to 3-D Endoanal Ultrasound and OASIS classification (each row shows the statistics for a single 10-fold cross-validation).

## Notes

### Competing Interest Statement

K.B. is a founder and management board member at OASIS Diagnostics, an author of the related patent and R&D strategy, independent consultant, and trainer of Takeda.
M.M., M.R., K.K., and P.I. are staff of OASIS Diagnostics. 
A.S. is an independent consultant of Ethicon, Takeda, Pfizer, Sofar. 
M.UM. is an independent consultant of Regen Lab.
H.H., P.J., M.UM., E.D., and E.GD. received remuneration as a study investigator.
H.H., C.R., and A.S. are independent consultants and members of the OASIS Diagnostics' Scientific Advisory Board. 
Others declare no conflicts of interest. 

### Clinical Trial

NCT04903977

### Author Declarations

The study was approved by the ethics committees respective for each study site: on 19 March 2021 by Ethics Committee of the Institute for Maternal and Child Care (no. 1/19.03.2021), on 27 April 2021 by Ethics Committee for Research with Medicines of the Health Areas of Leon and Bierzo (no. 2186), on 9 June 2021 by Ethics Committee of the University Hospital of Brno (no. 47/21Zdrav.), on 14 October 2021 by Ethics Committee at the Regional Medical Chamber in Warsaw (no. KB/1362/21) and on 25 July 2022 by Ethics Committee at AGEL Hospital Kosice-Saca (no. ONIRY 3/2/2020).

### Summary of Updates

This version of the manuscript has been revised to increase the readability and clarify the study design and aim.

