## Supplementary File S7 for "Detection of obstetric anal sphincter injuries using machine learning-assisted impedance spectroscopy: a prospective, comparative, multicentre clinical study"

**Supplementary File S7.** Performance metrics of the ONIRY system using the model trained based on the limited clinical study dataset (discrepant measurements, considered based on the arbitrary criterion set in Statistical Analysis Plan, were ignored), in the assessment relative to 3-D Endoanal Ultrasound and OASIS classification (each row shows the statistics for a single 10-fold cross-validation).

| **Seed** | **Accuracy [%]** | **Sensitivity [%]** | **Specificity [%]** | **F1** | **MCC** |
| --- | --- | --- | --- | --- | --- |
| 1 | 89.6 | 88.0 | 90.8 | 0.88 | 0.79 |
| 2 | 90.3 | 89.9 | 90.7 | 0.89 | 0.81 |
| 3 | 90.0 | 90.7 | 89.5 | 0.88 | 0.80 |
| 4 | 90.7 | 92.2 | 89.4 | 0.89 | 0.82 |
| 5 | 90.0 | 88.8 | 90.7 | 0.88 | 0.80 |
| 6 | 89.6 | 89.8 | 89.5 | 0.88 | 0.79 |
| 7 | 89.6 | 90.7 | 89.0 | 0.88 | 0.80 |
| 8 | 90.7 | 90.8 | 90.7 | 0.89 | 0.81 |
| 9 | 89.6 | 89.7 | 89.4 | 0.88 | 0.79 |
| 10 | 90.0 | 89.8 | 90.2 | 0.88 | 0.80 |
| **Mean ± SD** | **90.0 ± 0.4** | **90.0 ± 1.2** | **90.0 ± 0.7** | **0.88 ± 0.01** | **0.80 ± 0.01** |

- MCC = Matthew’s Correlation Coefficient; SD = Standard Deviation
