## Supplementary File S6 for "Detection of obstetric anal sphincter injuries using machine learning-assisted impedance spectroscopy: a prospective, comparative, multicentre clinical study"

**Supplementary File S6.** Performance metrics of the ONIRY system using the models trained based on all correct measurements from the clinical study, in the assessment relative to 3-D Endoanal Ultrasound and OASIS classification (each row shows the statistics for a single 10-fold cross-validation).

| **Seed** | **Accuracy [%]** | **Sensitivity [%]** | **Specificity [%]** | **F1** | **MCC** |
| --- | --- | --- | --- | --- | --- |
| 1 | 87.2 | 90.9 | 84.6 | 0.85 | 0.76 |
| 2 | 86.6 | 92.6 | 82.3 | 0.85 | 0.74 |
| 3 | 87.9 | 86.7 | 88.6 | 0.85 | 0.76 |
| 4 | 86.9 | 87.7 | 86.4 | 0.85 | 0.74 |
| 5 | 87.6 | 92.7 | 84.2 | 0.86 | 0.76 |
| 6 | 87.6 | 89.4 | 86.2 | 0.86 | 0.75 |
| 7 | 86.9 | 91.9 | 83.5 | 0.85 | 0.75 |
| 8 | 86.2 | 91.0 | 82.9 | 0.85 | 0.73 |
| 9 | 86.9 | 91.9 | 83.5 | 0.85 | 0.74 |
| 10 | 86.6 | 91.2 | 83.6 | 0.85 | 0.74 |
| **Mean ± SD** | **87.0 ± 0.5** | **90.6 ± 2.0** | **84.6 ± 1.9** | **0.85 ± 0.004** | **0.75 ± 0.01** |

MCC = Matthew’s Correlation Coefficient; SD = Standard Deviation
