## Supplementary File S5 for "Detection of obstetric anal sphincter injuries using machine learning-assisted impedance spectroscopy: a prospective, comparative, multicentre clinical study"

**Supplementary File S5.** The quality requirements for the impedance measurements.

It was decided, the impedance data from the measurement run will be excluded from further analysis if one or more of the following conditions are satisfied (suggesting the examination was performed incorrectly, even with a probe being outside of the anal canal):

- impedance value for any frequency is greater than 5000 Ohm
- phase shift for any frequency is greater than 0° (suggesting an inductive component of the impedance)
- resistance is over a certain arbitrary level for frequencies over 45 kHz
- reactance or resistance is under 0 Ohm for any frequency.
