## Supplementary File S4 for "Detection of obstetric anal sphincter injuries using machine learning-assisted impedance spectroscopy: a prospective, comparative, multicentre clinical study"

**Supplementary File S4.** Performance metrics of the ONIRY model trained based on data from the previous (pilot) clinical studies, depending on the heuristics chosen, with the 3-D Endoanal Ultrasound and the OASIS classification (>2 vs. ≤2) used as the reference method.

| **Heuristic** | **Accuracy [%]** | **Sensitivity [%]** | **Specificity [%]** | **F1 score** | **MCC** |
| --- | --- | --- | --- | --- | --- |
| H1 | 56.5 | 71.7 | 46.0 | 0.57 | 0.18 |
| H2 | 61.2 | 66.7 | 57.5 | 0.58 | 0.24 |
| H3 | 62.6 | 60.0 | 64.4 | 0.57 | 0.24 |
| H4 | 63.3 | 55.0 | 69.0 | 0.55 | 0.24 |
| H5 | 62.6 | 48.3 | 72.4 | 0.51 | 0.21 |
| H6 | 63.3 | 38.3 | 80.5 | 0.46 | 0.21 |

MCC = Matthew’s Correlation Coefficient
