## Supplementary File S3 for "Detection of obstetric anal sphincter injuries using machine learning-assisted impedance spectroscopy: a prospective, comparative, multicentre clinical study"

**Supplementary File S3.** Details of the *in silico* analyses conducted within Part II of the study.

1. Exploratory data analysis: A preliminary visual analysis of the data including impedance, phase shift, resistance, and reactance curves versus frequency of the application current; each measurement was visually inspected by the data scientist in the context of the presence or absence of OASI (per the reference diagnostic method), hereafter referred to as “labels”. If a measurement was highly outlying from the measurement range of the ONIRY system (did not correspond with the impedance in anal canal), it was excluded from the analysis.
2. Dimensionality reduction: Following calculation of the parameters describing the statistical features of the impedance, resistance, reactance, and phase shift curves, the feature selection and dimensionality reduction algorithms were used to determine the final set of parameters. This step aimed at simplifying the model and avoiding the use of irrelevant or redundant features for prediction, which could include bias in the ML modelling.
3. Modelling: Several modelling methods were applied. For this paper's purpose, only the results from the artificial neural networks method are presented. As applying this method resulted in the highest diagnostic performance metrics of ONIRY, it has been selected for the final model within the ONIRY system.
4. Ultimate performance analysis: Ten-fold cross-validations were performed, where metrics were calculated for each test fold and the average of these metrics was taken as the result for a given validation. As to obtain even more robust results, the process was repeated 10 times with different random seeds and the mean accuracies, sensitivities, and specificities were calculated for these 10-time 10-fold cross-validation processes. Training datasets were augmented for learning purposes. The metrics values obtained following this performance analysis were considered the final diagnostic performance measures of the ONIRY system.  The diagram illustrating the process of obtaining the final metrics is shown in Figure 1.

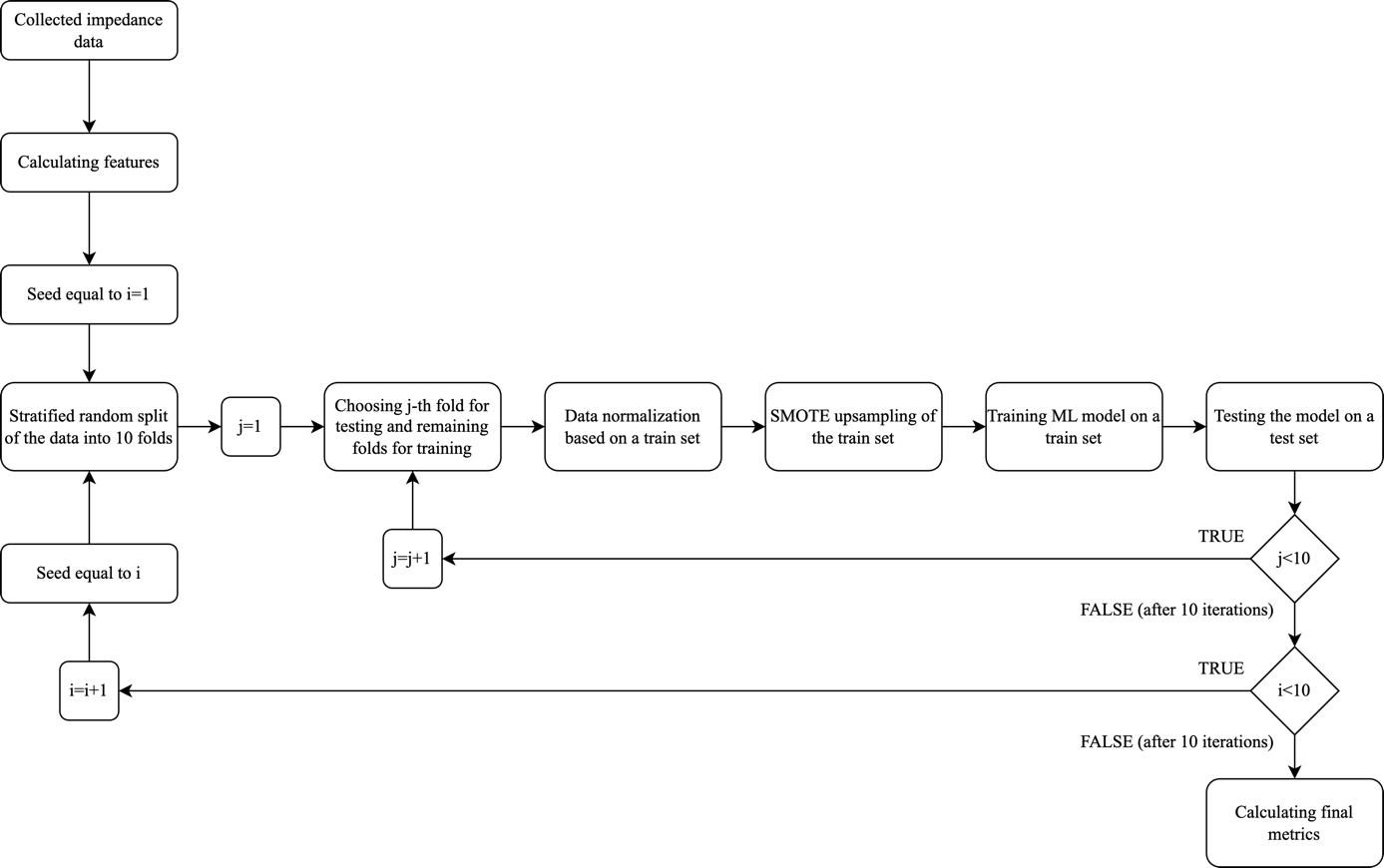

Figure 1. Diagram presenting the process of obtaining the final metrics in the *in silico* analyses.
