## Supplementary File S2 for "Detection of obstetric anal sphincter injuries using machine learning-assisted impedance spectroscopy: a prospective, comparative, multicentre clinical study"

**Supplementary File S2.** The description of the heuristics used.

The algorithm to interpret the ONIRY examination results was constructed based on data from the two previously completed pilot clinical studies. However, as this algorithm took parameters from the earlier versions of the anal probe (providing a single impedance measuring angle as the input), with regards to the current ONIRY Probe providing for 6 measuring angles, for Part I of the study, it was decided to test the performance of the ONIRY System based on different heuristics. The heuristic in which at least 2 out of 6 electrode angles must be labelled by the model as “injured” to classify the participant as “injured” (referred to as H2), was chosen for the ONIRY system used by study investigators during the clinical conduct (Part I of the study).

The heuristics were defined per the minimum number of measuring angles providing the “Injured” labels translating to the final “Injured” label. Thus, several analyses were performed for:

- Heuristic 1 (H1): at least one of six angles must be labelled by the model as “Injured” to classify the subject as “Injured”
- Heuristic 2 (H2): at least two of six angles must be labelled by the model as “Injured” to classify the subject as “Injured” (this heuristic was chosen to be implemented in the ONIRY System used by investigators during the clinical conduct)
- Heuristic 3 (H3): at least three of six angles must be labelled by the model as “Injured” to classify the subject as “Injured”
- Heuristic 4 (H4): at least four of six angles must be labelled by the model as “Injured” to classify the subject as “Injured”
- Heuristic 5 (H5): at least five of six angles must be labelled by the model as “Injured” to classify the subject as “Injured”
- Heuristic 6 (H6): all angles must be labelled by the model as “Injured” to classify the subject as “Injured”
