## Supplementary File S1 for "Detection of obstetric anal sphincter injuries using machine learning-assisted impedance spectroscopy: a prospective, comparative, multicentre clinical study"

**Supplementary File S1**. Schedule of assessments of the pivotal study (Part I – clinical conduct) valid at the time of the study completion (following Amendment no. 2 of the study protocol).

| **Study visit/days** | **V1**  **Day 1** | **V2**  **0-7 days post V1^a,b^** | **V3**  **1-28 days post V2** |
| --- | --- | --- | --- |
| Informed consent | X |  |  |
| Inclusion/exclusion criteria (see below) | X |  |  |
| Risk factors of obstetric anal sphincter injury | X |  |  |
| Medical history | X |  |  |
| History of pregnancies and deliveries | X |  |  |
| Wexner score^c^ | X |  |  |
| Body temperature | X | X^d^ | X |
| General physical examination | X | X^d^ | X |
| Gynecological examination | X |  |  |
| Assessment of anorectal area (proctological examination) | X |  |  |
| Body weight and height | X |  |  |
| Three-Dimensional Endoanal Ultrasound | X^e^ |  |  |
| Pregnancy test^f^ | X |  |  |
| Assignment to the appropriate study group | X^e^ |  |  |
| Impedance spectroscopy |  | X |  |
| Laboratory tests | | | |
| Blood tests (hematology, sodium, potassium, creatinine,  urea, C-reactive Protein, glucose (fasting or random) |  | X |  |
| Fecal calprotectin level |  | X |  |
| High-resolution anorectal manometry |  |  | X^g^ |
| Vital signs | X | X^d^ | X^g^ |
| Electrocardiography | X | X |  |
| Adverse event recording | X | X | X |

^a^If the endoanal ultrasound and study group allocation were performed on the next day following V1, the V2 could be scheduled on this day at the earliest

^b^If OASI was diagnosed at V1, and a prompt primary surgical repair was decided, the V2 visit must have been conducted prior to this repair, unless deemed unsafe for the participant

^c^Assessable if V1 visit conducted ≥3 days after delivery

^d^Not performed in case the V1 and V2 visits were conducted on the same day

^e^Could be performed up to 1 day following the V1 visit

^f^Only for participants recruited ≥4 weeks after delivery with no lactation at V1 visit; however, participants with a positive pregnancy test may have been enrolled as long as gynecologic ultrasound performed at V1 visit showed no signs of a new pregnancy, no sings suggestive of placental tissue remaining in utero, or other abnormality of the uterus

^g^In case no anorectal manometry was performed in the given subject, the V3 visit could be conducted by telephone as to only monitor the adverse events

**Inclusion criteria**

The study group-specific inclusion criteria were set. For Group A, there were: no clinical signs of any degree perineal tear, no clinical signs or symptoms of any damage involving anal sphincters, presence of not more than 1 of the following OASIs risk factors related to the last delivery: prolonged second phase of delivery, fetal shoulder dystocia, birth weight of the neonate >4 kg, induction of delivery using oxytocin, or head circumference of the neonate ≥34 cm. For Group B, it was clinically confirmed 1st or 2nd degree perineal tear according to the OASIS classification (including episiotomy and uncontrolled crotch rupture) occurred at the last delivery. Similarly, for Group C, it was clinically identified 3rd or 4th degree perineal tear according to the OASIS classification (injury involving anal sphincters) that occurred at the last delivery (regardless of primary repair).

**Exclusion criteria**

The exclusion criteria were: any acute, uncontrolled disease (except for hemorrhoidal disease), chronic diseases not treated or not stable on treatment, symptoms of FI due to a disease other than diagnosed or suspected OASI, previous surgery for OASI (primary or secondary), FI or anal prolapse, except for a primary repair of anal sphincter damage performed after the last delivery (allowed), any surgery in perineal or rectal area, including surgery for OASI, planned for the study period, presence of inflammatory bowel diseases during exacerbation phase, any treatment during last 12 months for severe, progressive, uncontrolled cardiological, pulmonary, nephrology, contagious or psychiatric illness that could increase subject's risk due to participation in the study, disease other than OASI so far undiagnosed and reported during the first visit (V1) or within 7 days prior to it, present or suspected malignancy or previous oncological treatment in the last 5 years, implanted cardiac stimulator or cardioverter-defibrillator, clinically significant cardiac arrhythmias observed in ECG examination or reported in history for the last 12 months, fever (>37.5°C) at enrolment, history of major surgery in perineal or rectal area (other than for OASI) or severe trauma of perineum or rectum, use, or need for use, of an anal suppository or other anally administered drug, or cosmetic for the perianal area, within 12 hours prior to impedance spectroscopy examination, a positive pregnancy test (only for subjects recruited ≥4 weeks after delivery with no lactation at V1 visit; however, subjects with a positive pregnancy test could have been enrolled as long as gynecologic ultrasound performed at V1 visit shows no signs of a new pregnancy, no sings suggestive of placental tissue remaining in utero, or other abnormality of the uterus).
